# 5-hydroxymethylcytosine sequencing in plasma cell-free DNA identifies unique epigenomic features in prostate cancer patients resistant to androgen deprivation therapies

**DOI:** 10.1101/2023.10.13.23296758

**Authors:** Qianxia Li, Chiang-Ching Huang, Shane Huang, Yijun Tian, Jinyong Huang, Amirreza Bitaraf, Xiaowei Dong, Marja T. Nevalanen, Manishkumar Patel, Jodie Wong, Jingsong Zhang, Brandon J. Manley, Jong Y. Park, Manish Kohli, Elizabeth M. Gore, Deepak Kilari, Liang Wang

## Abstract

**Background:** Currently, no biomarkers are available to identify resistance to androgen-deprivation therapies (ADT) in men with hormone-naive prostate cancer. Since 5-hydroxymethylcytosines (5hmC) in gene body are associated with gene activation, in this study, we evaluated whether 5hmC signatures in cell-free DNA (cfDNA) predicts early resistance to ADT.

**Results:** We collected a total of 139 serial plasma samples from 55 prostate cancer patients receiving ADT at three time points including baseline (prior to initiating ADT, N=55), 3-month (after initiating ADT, N=55), and disease progression (N=15) within 24 months or 24-month if no progression was detected (N=14). To quantify 5hmC abundance across the genome, we used selective chemical labeling sequencing and mapped sequence reads to individual genes. Differential methylation analysis in baseline samples identified significant 5hmC difference in 1,642 of 23,433 genes between patients with and without progression (false discovery rate, FDR<0.1). Patients with disease progression showed significant 5hmC enrichments in multiple hallmark gene sets with androgen responses as top enriched gene set (FDR=1.19E-13). Interestingly, this enrichment was driven by a subgroup of patients featuring a significant 5hmC hypermethylation in the gene sets involving *AR*, *FOXA1* and *GRHL2*. To quantify overall activities of these gene sets, we developed a gene set activity scoring algorithm and observed significant association of high activity scores with poor progression-free survival (P<0.05). Longitudinal analysis showed that the high activity scores were significantly reduced after 3-months of initiating ADT (P<0.0001) but returned to higher levels when the disease was progressed (P<0.05).

**Conclusions:** This study demonstrates that 5hmC-based activity scores from gene sets involved in *AR*, *FOXA1* and *GRHL2* may be used as biomarkers to determine early treatment resistance, monitor disease progression, and potentially identify patients who would benefit from upfront treatment intensification.

## Background

Prostate cancer is the most common cancer in men and the second leading cause of cancer-related death(1). Prostate cancer cells are reliant on circulating androgens to activate endogenous androgen receptor (*AR*)(2). Suppression of testicular androgens by castration (medical or surgical) is the mainstay of treatment for metastatic disease(3). Androgen deprivation therapies (ADT) achieves a remission in 80–90% of men with advanced prostate cancer as measured by monitoring a serum prostate specific antigen (PSA) decrease and an average progression-free interval of 12–33 months(4). Despite the efficacy of ADT, disease progression from a hormone-sensitive state is inevitable to a castration-resistant state(5). To address this issue, the therapeutic strategy has evolved with the addition of other systemic agents. The combination of ADT with docetaxel or with new androgen receptor signaling inhibitors (ARSI) has shown substantial benefits (6). Because docetaxel and ARSI have different mechanisms of action on androgen signaling and prostate cancer cells, the combination may enhance the treatment effect(7–9). However, patients with similar clinical pathological factors may respond differently, suggesting phenotypic heterogeneity and potential role of genetic background in the treatment response. To offer more effective treatment, it will be essential to understand which patient group would truly benefit from this therapy. Development of biomarkers will facilitate the selection of the most appropriate treatment.

Currently, tissue biopsies have demonstrated a limited role outside of histologic diagnosis for treatment decision and are impractical to perform routinely in clinical practice due to the bone-predominant metastasis of prostate cancer. To address this challenge, recent research focus has turned to the development of minimally-invasive biomarkers from body fluids (10). Blood serological biomarker PSA is commonly used for screening and diagnosis of prostate cancer, as well as for monitoring treatment response (11). However, PSA does not predict systemic treatment response. In search for more predictive biomarkers, cell-free DNA (cfDNA) circulating in blood has attracted significant attention. The minimally invasive detection of somatic variations using blood samples offers substantial advantages over tissue biopsy as it can detect entire genetic makeup from tumor tissues. The easily accessible nature of blood makes it an ideal sample source for real-time and dynamic monitor of the treatment response and disease progression(12).

DNA methylation at specific genomic regions is characteristic of human cancers and has been used as a specific biomarker for cancer detection and clinical outcome prediction (13). In addition to commonly reported 5-methylcytosines in the genome, 5-hydroxymethylcytosines (5hmC) are also abundant. Recent genome-wide sequencing maps of 5hmC in various mammalian cells and tissues support its regulatory role for gene expression (14). The 5hmC is enriched at transcriptionally active regions (such as gene bodies) and may represent dynamically activated transcription rather than constitutively expressed house-keeping genes (15). Therefore, 5hmC has emerged as a novel class of cancer epigenomic biomarkers, with a recent study showing cfDNA 5hmC as a prognostic factor in metastatic prostate cancer (16). So far, however, little is known about the potency and reliability of the cell-free 5hmC as a predictive biomarker of treatment response for metastatic hormone-sensitive prostate cancer under standard of care ADT combination therapies. To address this question, we employed the 5hmC-Seal (17), a highly sensitive and selective chemical labelling-based sequencing technology, to profile the full spectrum of 5hmC in the hormone-sensitive patients with three serial time points of blood collection from pre-ADT to 3 months post-ADT and at the time of progression. Our results reveal that 5hmC-based epigenomic features are associated with treatment response and classify early resistance to standard of care ADT.

## Patients and Methods

### Patient cohort

A total of 55 prostate cancer patients were enrolled in the prospective cohort study. Among those, 30 patients were recruited from the Medical College of Wisconsin and additional 25 patients were recruited from the Milwaukee VA Medical Center, United States. All patients were diagnosed with advanced prostate cancer and met the NCCN treatment guideline for ADT-based treatments. Blood was collected before initiating ADT (baseline), 3 months after initiating ADT and at 24 months after ADT or at disease progression within the 2-year follow-up. Written informed consent was obtained from all participants prior to enrollment. Progression was defined as either radiological (as defined by Prostate Cancer Working Group 2 criteria)(18) or clinical (defined as worsening disease-related symptoms necessitating a change in anti-cancer therapy and/or deterioration in ECOG performance status ≥2 levels)(19). The study design and workflow are shown in **Figure 1**.

**Figure 1.**
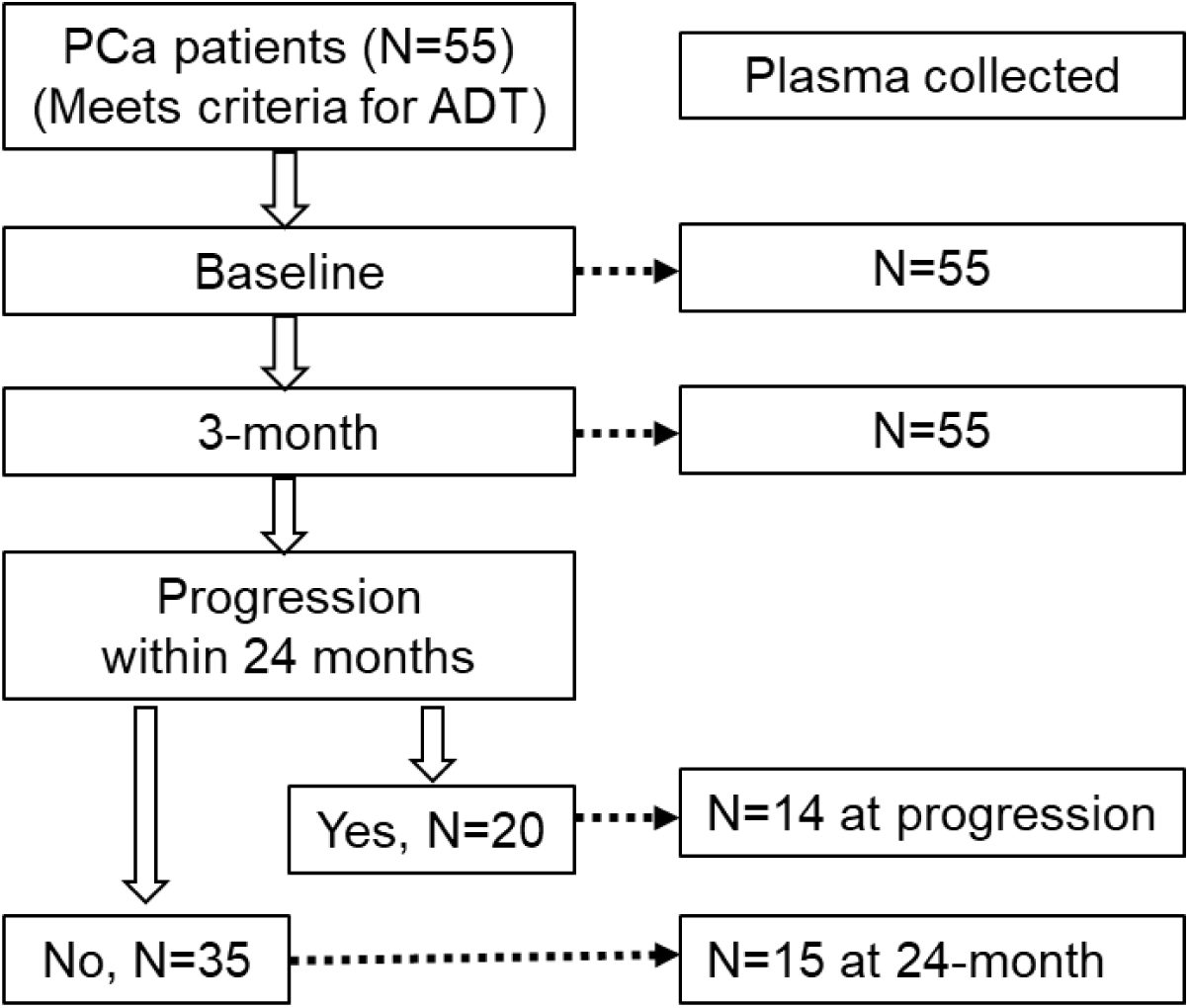
Study design and blood collection. A total of 55 patients who met the NCCN guidelines for ADT were enrolled. Blood was collected at baseline (N=55), 3-month (N=55), disease progression (N=14) and 24-month if no progression (N=15).

### Blood sample processing

Plasma was prepared within 2 hours after blood draw using EDTA tubes. Collected blood samples were first centrifuged at 1000g for 10 minutes at room temperature to separate plasma. The platelets-rich plasma was immediately centrifuged again at 5000g for additional 10 minutes to collect platelets-poor plasma before stored at -80°C. cfDNAs were extracted from 400-1000μl of platelets-poor plasma using QIAamp DNA Blood Mini Kit (Qiagen). Final DNA eluent (50μl) was quantified by a Qubit 2.0 Fluorometer (Life Technology) and stored at -80°C until use. Additionally, cfDNA from plasma samples and gDNA from peripheral blood mononuclear cells (PBMCs) in 8 healthy subjects were also collected.

### Spike-in controls for 5hmC enrichment

The 5hMe-Seal method has been previously published(17, 20). In brief, the spiked-in control was generated by PCR-amplified lambda DNA using a cocktail of dATP/dGTP/dTTP and one of the following: dCTP, dmCTP or 10% dhmCTP (Zymo)/90% dCTP. Primers sequences are as follows: dCTP FW-5′-CGTTTCCGTTCTTCTTCGTC-3′, RV-5′ TACTCGCACCGAAAATGTCA-3′; dmCTP FW-5′-GTGGCGGGTTATGATGAACT-3′, RV-5′-CATAAAATGCGGGGATTCAC-3′; 10% dhmCTP/90% dCTP FW-5′-TGAAAACGAAAGGGGATACG-3′, RV-5′-GTCCAGCTGGGAGTCGATAC-3′. The spike-in probes were utilized to monitor the robustness and sensitivity of 5hmC-Seal. In total, 0.2 million copies of 5hmC and no 5hmC spike-ins were first mixed and then added into cfDNA samples before library preparation. The input control samples omitted the 5hmC pull-down step to generate non-5hmC enrichment libraries. After sequencing, spike-in reads were called, and enrichment ratios were calculated.

### 5hmC library construction and high-throughput sequencing

5hmC libraries were constructed as described previously(20). Briefly, the 5-10ng cfDNA was ligated with sequencing adaptors. The ligated DNA was incubated in a 25ul reaction solution containing HEPES buffer (50 mM, pH 8.0), MgCl2 (25 mM), N3-UDP-Glc (100 mM, Active Motif), and 12.5 U T4 phage β-glucosyltransferase (βGT, Thermo Fisher) for 1 h at 37 °C. Then, 2.5µl DBCO-PEG4-biotin (20 mM stock in DMSO, Click Chemistry Tools) was directly added to the reaction mixture and incubated for 2 h at 37°C. Subsequently, the DNA Clean & Concentrator™-5 (ZYMO Research) was used to purify the DNA. Thereafter, the purified DNA was incubated with C1 streptavidin beads (5ul, Life Technologies) for 15 min at room temperature. The beads subsequently underwent eight 5-min washes with buffer (5mM Tris pH 7.5, 0.5 mM EDTA, 1 M NaCl and 0.1% Tween 20). All binding and washing were done at room temperature with gentle rotation. Beads were then resuspended in water and amplified with 12-15 (cfDNA) or 9 (whole blood genomic DNA) cycles of PCR amplification using KAPA Library Amplification Kit (Kapa Biosystems). A separate non-enrichment control library from cfDNA was also made by direct PCR amplification from ligated DNA without labeling and capture. The amplified products were purified using AMPure XP beads and used for high-throughput 75 cycle single end sequencing on the Illumina NextSeq 500 platform.

### Data processing and normalization

Pre-alignment quality control was performed for the raw sequenced reads using fastp (Version 0.20.1) (21) with the default settings. The raw 5hmC-Seal data were first cleaned for adaptor sequences, followed by aligning to the human genome (hg19)using Bowtie-2 (Version 2.4.2)(23). SAMtools (Version 1.11) command lines were used to convert the file format from SAM to BAM, followed by sorting, indexing, and duplicate read removing (24). The alignment excluded the ENCODE blacklisting regions (25). Enrichment regions were identified by model-based analysis of ChIP-Seq (MACS) (26). FeatureCounts from the Subread package (Release 2.0.3) were used to call read counts for each gene [27]. The Human Release 19 comprehensive gene annotation (GRCh37.p13) was used as a reference. The read counts for either 1,000bp bins or individual genes were extracted using bedtools (27) and further normalized using DESeq2 (28), which performs the variance-stabilizing transformation to correct for sequencing depth and library size.

### Calculation of gene set activity scores

Gene lists of selected gene sets were downloaded from three data sources. The hallmark androgen response gene set was derived from GSEA website. Lists of co-expressed genes with *AR*, *FOXA1*, and *GRHL2* as well as *AR* targets by ChIP-seq were downloaded from Enrichr gene set library (29). The list of androgen signaling genes was copied from the publication (30). To calculate gene set activity score, we transformed read counts into log2 ratio of read counts in each individual genes between each patient and a pooled healthy control. We then identified mean value of the log2 ratios in the gene set and further multiplied the mean value by 100. The formula is the following:

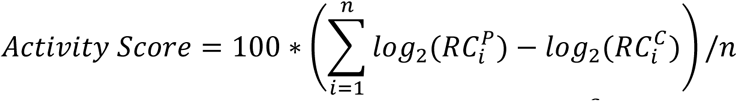

where 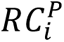 is the normalized read count of gene *i* for a patient, 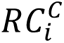 is the average normalized read count of gene *i* among the 7 healthy controls, and *n* is the total number of genes in a gene set.

### Statistical analyses

DESeq2 was utilized to identify the differentially methylated genes between progressed and nonprogressed patients, and among three epigenetic groups. To correct for multiple testing, a false discovery rate (FDR) was used, with FDR less than 0.1 being used as the cutoff. Student’s t-tests were used to compare the activity score, ctDNA fractions and PSA values among different groups of patients. Kaplan Meir analysis was used to identify epigenomic features that were associated with progression-free survival. Analysis of covariance (ANCOVA) was applied to adjust for the effect of ctDNA fraction and clinical covariates. ctDNA fraction was estimated by ichorCNA, a tool for estimating the fraction of tumor in cell-free DNA from ultra-low-pass whole genome sequencing (31). For differences in gene set activity scores, ctDNA fraction and PSA at baseline, a two-sided p value less than 0.05 was considered significant.

## Results

### Clinical characteristics and plasma collection

In the 55 hormone-naïve prostate cancer patients, the median age was 69 (range 49-94). At enrollment, 44 of the 55 patients showed bone or soft tissue metastasis and the remaining 11 presented with non-metastatic castrate status (biochemical recurrence). Among the patients with metastasis, 18 had high volume and 26 had low volume disease (CHAARTED criteria). During 24-month follow-up, 20 patients showed disease progression (defined as early resistance to ADT in this study) and 35 had no sign of disease progression. More clinical characteristics of this cohort are summarized in **Table 1** (**Additional Table S1** for detail). Plasma samples from baseline (prior to ADT) and 3-month post ADT were collected in all 55 patients. Additional plasma samples were collected either at the time of progression in 14 of 20 progressed patients or at 24-month post-ADT in 15 of 35 non-progressed patients.

**Table 1.**
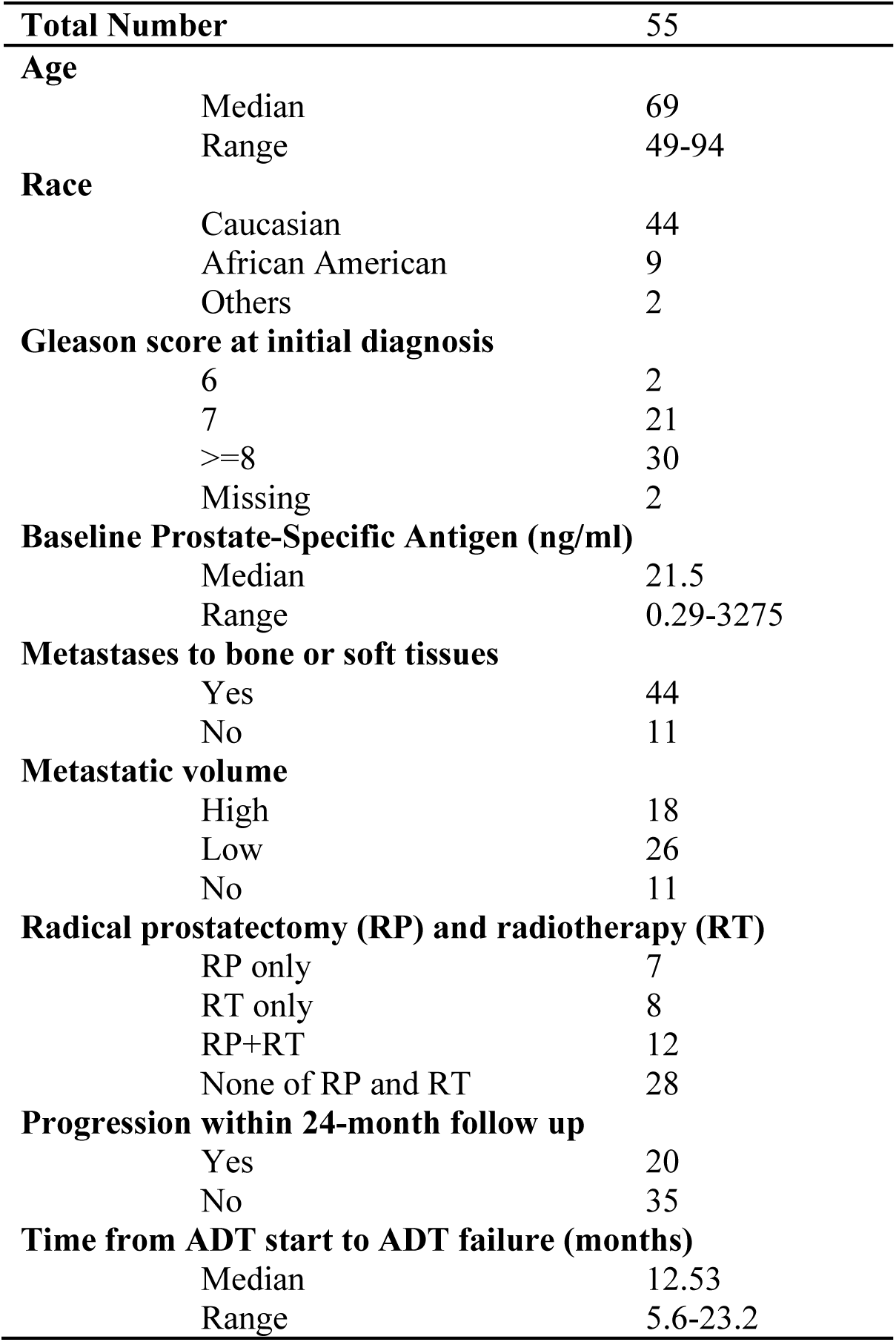
Clinical characteristics of patients.

### 5hmC sequencing in cfDNA shows high enrichment efficiency and specificity

To determine 5hmC enrichment efficiency and specificity, we spiked in a pool of 180 bp amplicons containing either C, 5mC or 5hmC into cfDNA during library preparation. We first performed PCR analysis in 5hmC-enriched libraries and observed amplicons in 5hmC-containing DNA only (**Additional Figure S1A**). This result was confirmed in the final sequencing libraries, which showed over 100-fold enrichment in reads mapping to 5hmC spike-in DNA (**Additional Figure S1B-C**). We also examined duplication rate in these 5hmC libraries. With a median read count of 18.3 (6.03-42.43) million/sample, the sequencing libraries showed 98% (95-99%) mappable reads and 75% (72-78%) unique (nonduplicate) reads (**Additional Table S2**).

### 5hmC profiles in cfDNA are different from PBMC-derived gDNA

To evaluate the distribution of 5hmC in cfDNA, we first performed enrichment analysis using 1kb window bins across the genome. Peak detection in healthy control cfDNAs showed that 66.7% of 5hmC-enriched regions were in the gene bodies including introns (47%) and CDS (19.7%), whereas only 6.1% enriched regions were found in 5’ UTRs (**Figure 2A**). We then compared gene body read counts between cfDNA from healthy controls and peripheral blood gDNA. From a total of 23,433 genes with median raw read count ≥8, we observed 11,688 differentially methylated genes (DMGs) including 5,819 hypermethylated genes and 5,869 hypomethylated genes in cfDNA (FDR < 0.1, **Figure 2B**). Enrichment analysis showed that the hypermethylated DMGs were enriched in apoptotic process and immune response (**Figure 2C**), whereas hypomethylated genes were enriched in cell structure, cell adhesion, and neuron protection (**Figure 2D**).

**Figure 2.**
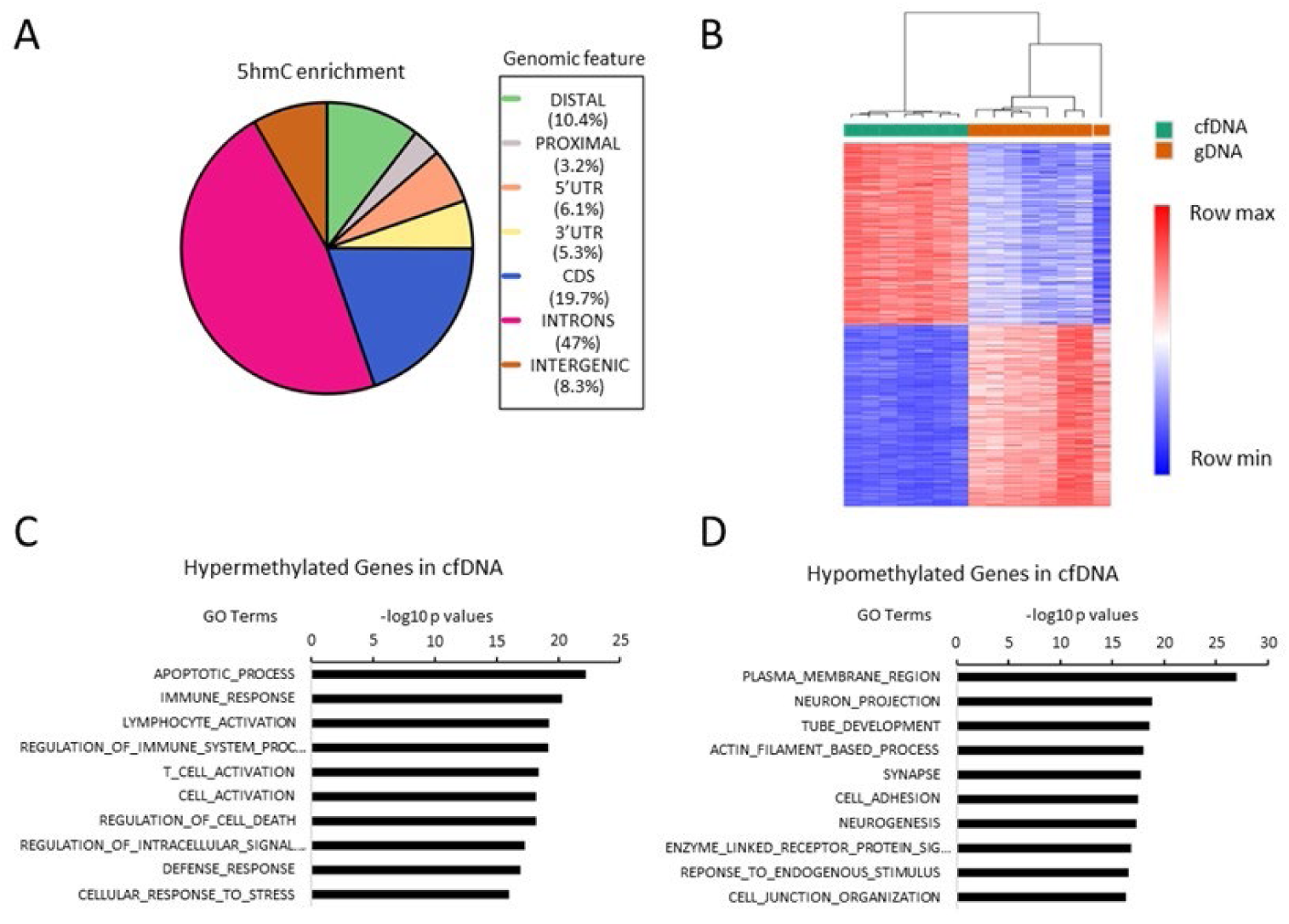
Distribution of 5hmC-enriched regions and genes. A. Genome distribution of 5hmC-enriched regions. B. Heatmap showing differentially methylated genes between cfDNA and blood mononuclear cells-derived gDNA. C. Hallmark gene set enrichment in cfDNA hypermethylated genes. D. Hallmark gene set enrichment in cfDNA hypomethylated genes.

### Baseline plasma cfDNAs show diverse 5hmC epigenomic profiles

To test 5hmC differences at baseline cfDNAs, we compared read count in each individual gene between patients with disease progression and those without progression during 24-month follow up. Among the 23,433 genes tested, we identified 1,642 DMGs (FDR < 0.1) including 1,008 hypermethylated and 634 hypomethylated genes in the patients with progression (**Additional Table S3)**. To evaluate molecular mechanisms underlining the differential methylation, we performed enrichment analysis using these DMGs in hallmark gene sets. This analysis showed significant enrichment in multiple gene sets with androgen response (FDR=1.19E-13) and estrogen response early (FDR=5.50E-13) as top 2 gene sets in hypermethylated genes, and complement (FDR=1.13E-3) and inflammatory response (FDR=2.93E-03) as top 2 gene sets in hypomethylated genes (**Figure 3A-B**). Interestingly, these DMGs revealed significant epigenomic heterogeneity within both clinical groups of patients. Hierarchical clustering analysis showed that a fraction of progressed patients demonstrated unique epigenomic features that separated them from all other patients although most progressed patients tended to cluster with non-progressed patients (**Figure 3C**). Specifically, although the 55 patients were clinically classified into two groups, they were epigenetically classified into three groups by unsupervised analysis. EpiGroup 1 included these patients with disease progression (N=6) and showed unique epigenomic features. EpiGroup 2 featured highly similar epigenomic profiles between the subgroups of progressed (N=14) and non-progressed patients (N=17). EpiGroup 3 was a unique cluster of patients without progression (N=18). The three epigroups demonstrated significant differences in progression-free survival (**Figure 3D**).

**Figure 3.**
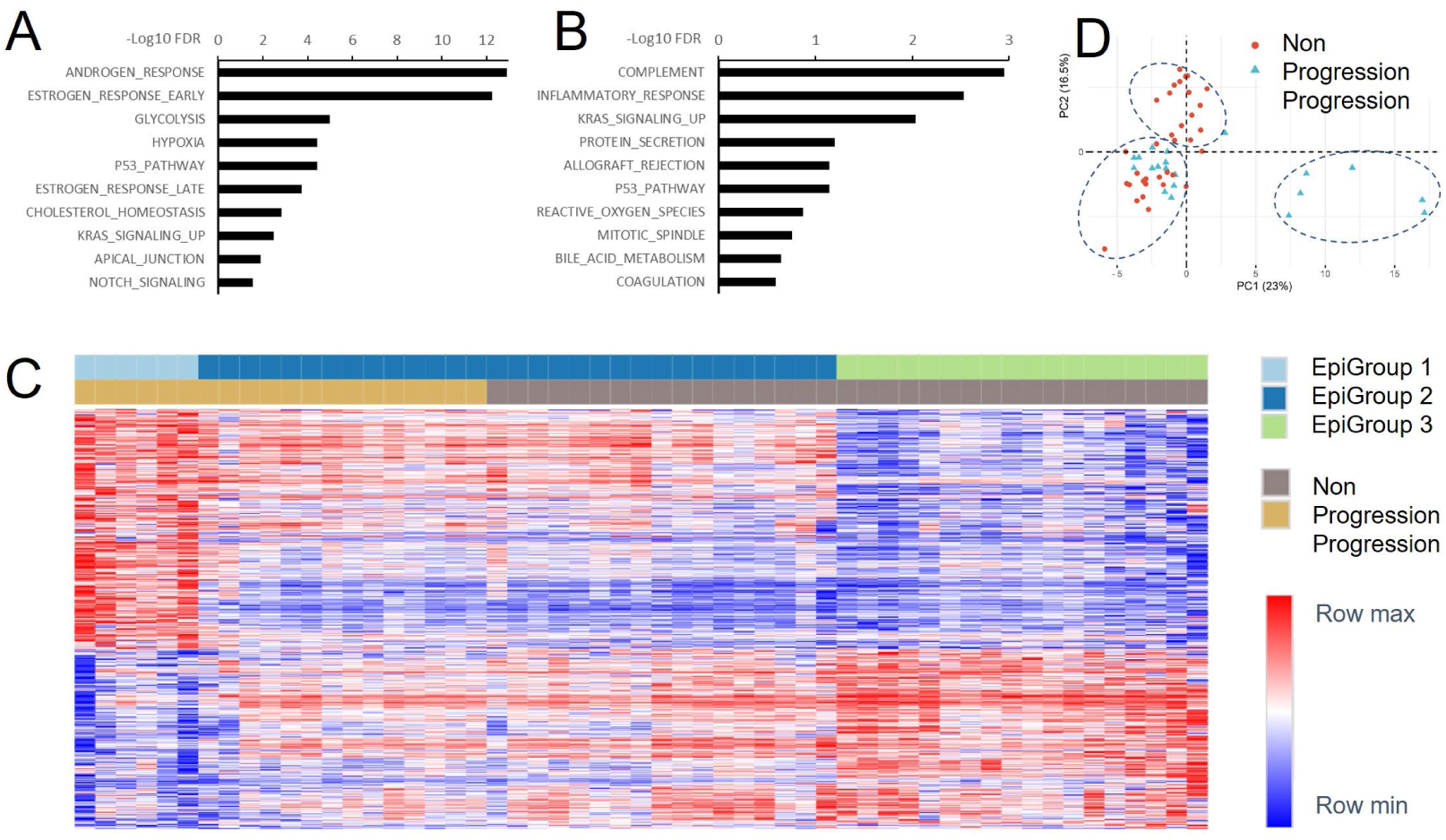
Gene set enrichment and epigenomic heterogeneity. A. Gene set enrichment in hypermethylated genes of baseline samples when comparing progressed to non-progressed patients. B. Gene set enrichment in hypomethylated genes of baseline samples when comparing progressed to non-progressed patients. C. Heatmap showing distinct epigenomic groups in both progressed and non-progressed patients. D. Kaplan-Meir analysis showing survival differences in three epigroups.

### Patients with different epigenomic features show 5hmC hypermethylation in unique signaling pathways

To evaluate molecular mechanisms of the epigenomic heterogeneity, we compared the 5hmC profiles using the 23,433 genes among three EpiGroups. This comparison identified 13,220 (EpiGroups 1 vs. 2), 5,046 (EpiGroups 1 vs. 3) and 12,603 (EpiGroups 2 vs. 3) DMGs (**Additional Table S4**). Gene enrichment analysis showed significant differences in a wide variety of signaling and regulatory pathways. Notably, when compared to either EpiGroup 2 or EpiGroup 3, the EpiGroup 1 consistently showed that the androgen response was the most significantly hypermethylated gene set (FDR≤5.63E-10), while immune responses (complement, allograft rejection and inflammation) were among the top hypomethylated gene sets (FDR≤ 1.04E-4) (**Additional Figure S2A-D, Table S5**). Of note, these hyper- and hypo-methylated gene sets had a similar trend to the significant gene sets when comparing two clinical groups (**Figure 3A-B**). Clearly, EpiGroup 1 is the main driver of epigenomic differences between patients with progression and patients without progression. This unique subgroup is characterized by 5hmC hypermethylation (activation) of the androgen response gene set, and 5hmC hypomethylation (inactivation) of immune responses. When comparing Epigroups 2 with 3, we observed significant enrichment in hallmark P53 pathway and mitotic spindle gene sets (FDR≤ 2.13E-6) with an increased methylation in EpiGroup 2 (**Additional Table S5**)

### Gene set activity scores of baseline samples classify patients with different clinical outcomes

To quantify the active status of these gene sets, we developed a gene set activity scoring algorithm using a mean value of log2 ratios among all genes of selected gene sets. We first calculated the activity score using 97 genes that overlapped with hallmark androgen response gene set. The average activity scores were statistically different (p=5.43E-05) between patients with progression (score= 6.62 ± 11.65) and patients without progression (score= -3.23 ± 4.90) (**Figure 4A**). Subgroup analysis showed that the difference was driven by EpiGroup 1. Specifically, the average activity scores were 22.27, -0.84 and -4.90 in EpiGroups 1, 2 and 3, respectively. The activity score was significantly higher in EpiGroup 1 than in EpiGroup 2 (p=4.73E-14) and EpiGroup 3 (p=1.73E-08) (**Figure 4B**). Clearly, the patients in EpiGroup 1 had the highest activity scores that were distinguished from the remaining patients regardless of progression status (**Figure 4C**). Meanwhile, to evaluate whether the activity score was predictive of disease outcome, we performed Kaplan-Meier analysis and observed significant association of the activity score with progression-free survival (p=0.0006, HR= 5.35, **Figure 4D**). Specifically, comparing to patients with high activity score (median value as cutoff), the patients with low-activity score showed approximately 43% progression-free survival benefit after 24 months of ADT-related treatment.

**Figure 4.**
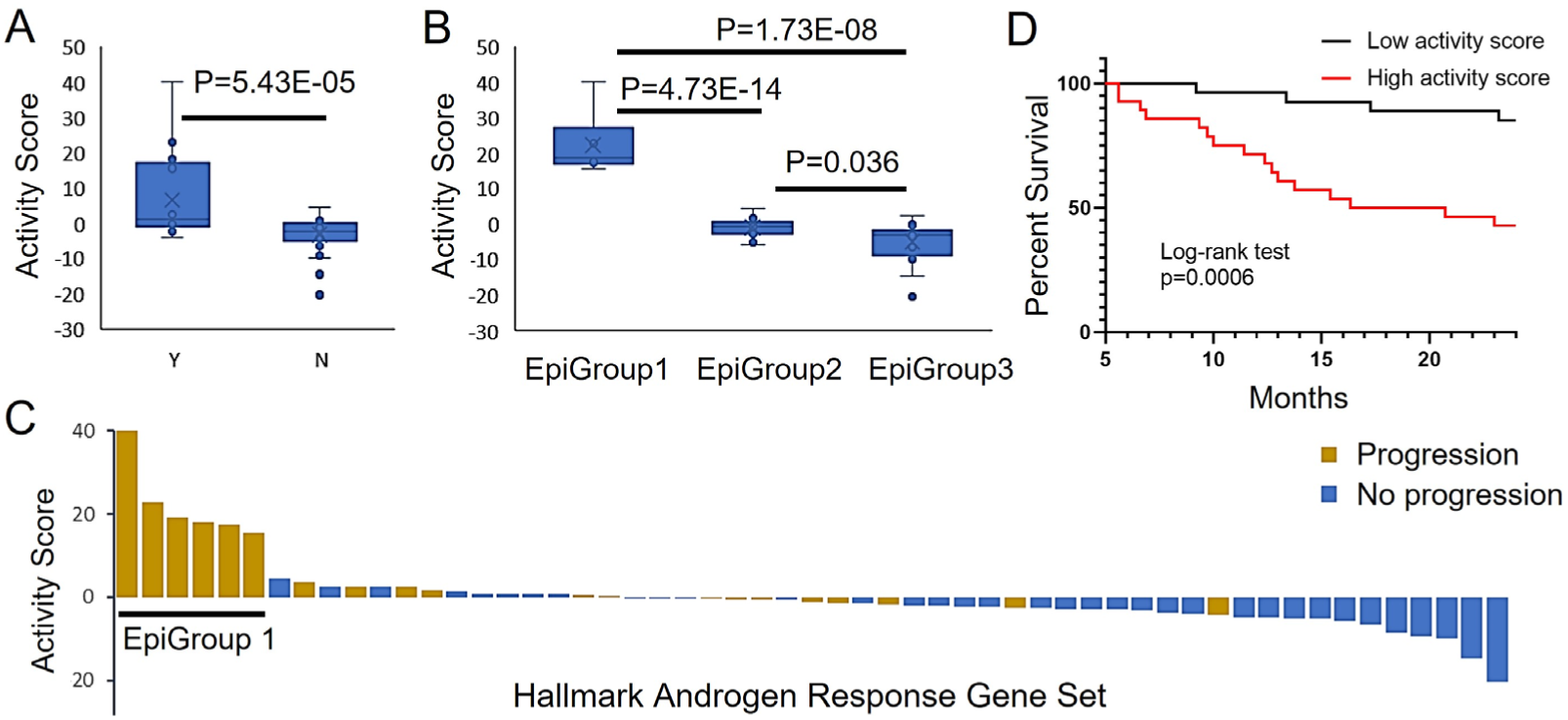
Activity scores in androgen signaling gene sets separate patients with different clinical outcomes. A. Activity score of hallmark androgen response gene set is significant higher in patients with progression (Y) than without progression (N). B. Activity score in EpiGroup 1 is significantly higher than EpiGroups 2 and 3. C. Waterfall plot shows clear-cut activity score difference in EpiGroup 1. D. High activity score is associated with poor progression-free survival.

In addition to hallmark androgen response gene set, we also tested other androgen-related gene sets including *AR* target genes (ChIP-seq), *GRHL2*-coexpressed genes, *FOXA1*-coexpressed genes, *AR*-coexpressed genes (29) and *AR* signaling pathway (30). Overall distribution of these activity scores in the selected gene sets are shown in **Figure 5A**. To test the activity differences, we first compared two groups of patients with different clinical outcomes in baseline samples. This comparison showed significantly higher activity scores in patients with progression than those without progression (**Figure 5B**). We then compared three groups of patients with different epigenomic features. This analysis showed consistently higher activity scores in EpiGroup 1 than in EpiGroups 2 and 3 patients (**Figure 5C**). In addition to hallmark androgen response gene set (**Figure 4B**), 4 of other 5 androgen-related gene sets (*GRHL2*-, *FOXA1*- and *AR*-coexpressed gene sets, and androgen signaling gene set) also showed higher activity score in EpiGroup 2 than in EpiGroup 3 (**Figure 5C**). Kaplan-Meier analysis demonstrated poor progression-free survival in patients with high activity score (**Figure 5D**).

**Figure 5.**
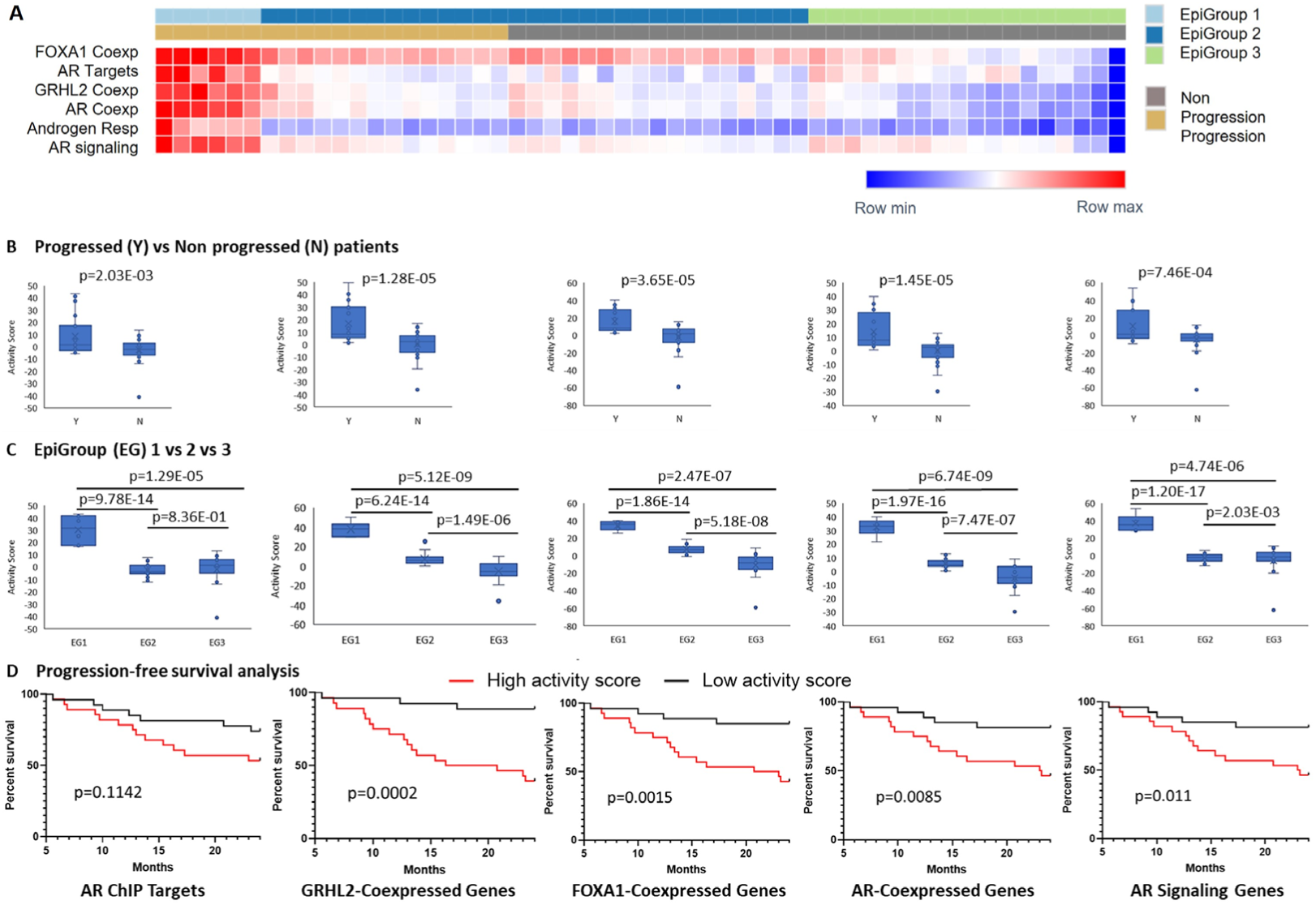
Activity scores in patients with different clinical outcomes or among patients with different epigenetic status. A. Heatmap showing overall distribution of activity scores in androgen response gene sets. B. Significant activity score differences between progressed (Y) and non-progressed (N) patients. C. Significant activity score differences among three EpiGroups (EG1, EG2, EG3). D. Association of high activity scores with poor progression-free survival.

### Baseline gene set activity score predicts treatment resistance independent of clinical factors and androgen receptor amplification

Since multiple clinical factors have shown a potential association with disease progression, we tested whether these clinical factors had any effect on discriminative performance of the gene set activity scores. We first tested Gleason score at diagnosis, age at enrollment, metastatic volume, and baseline PSA for their potential association with disease progression in the 55 patients. Among all clinical factors tested, only PSA level at enrollment showed significant association with disease progression (p=2.35E-05) (**Additional Figure S3A**). Because the whole genome 5hmC sequencing data can be used for estimating ctDNA fraction (20), we then calculated the ctDNA percentage using a previously published algorithm(32). We found that the higher baseline ctDNA percentage was also associated with disease progression (p=0.0035) **(Additional Figure S3B**). Because of their clinical associations, we further performed an analysis of covariance to evaluate the effect of these factors on the association of activity scores with clinical outcomes. By applying baseline PSA level and ctDNA percentage as covariates, our analysis showed that baseline activity scores in patients with disease progression were still significantly higher than the ones without progression in 4 of six gene sets (**Additional Table S6**).

Additionally, we also evaluated 5hmC status at *AR* locus. We plotted the mean values of read count log2 ratios in the genes flanking *AR* and observed significantly higher log2 ratios in EpiGroup 1 than EpiGroups 2 and 3 at *AR* locus. However, flanking gene loci did not show significantly high read count in the EpiGroup 1 (**Additional Figure S3C**). Since the 5hmC capture assay is highly specific, the high level of 5hmC at *AR* but not nearby gene loci indicates specific activation of *AR* expression. Additionally, the *AR* is commonly co-amplified with nearby genes (32, 33), the lack of co-amplification also supports the notion that the activation of androgen response gene sets may not be caused by *AR* amplification in this study cohort.

### Pharmacodynamics of gene set activity scores reflects treatment response and serves as surrogates to monitor disease progression

To evaluate whether 5hmC profile changes reflect treatment outcomes, we compared pharmacodynamics of the entire 23,433 genes at different blood collection times in three groups of patients with different epigenomic features. Although patients in EpiGroup 2 did not show significant difference between different timepoints, we did observe significant changes when comparing 3-month to baselines in EpiGroup 1 and EpiGroup 3. Specifically, 3-month treatment caused significant 5hmC changes in 4,315 genes including 2,627 reduced and 1,688 increased methylations in EpiGroup 1, and 4,112 genes including 1,803 reduced and 2,309 increased methylation in EpiGroup 3 (FDR<0.1, **Additional Table S7**). Interestingly, enrichment analysis in EpiGroup 1 showed the most significant methylation reduction in gene sets of androgen and estrogen responses while the most significant methylation increases in the gene sets of immune responses (**Additional Table S8**). Similarly, enrichment analysis in EpiGroup 3 showed that 3-month treatment induced significant methylation increases in gene sets of mitotic spindle and p53 pathway (**Additional Table S9)** although we did not observe significant methylation reduction in any hallmark gene sets.

We also compared the pharmacodynamics of gene set activity scores at three blood collection times. When compared to baseline, 3-month treatment in EpiGroup 1 patients significantly reduced activity scores in all androgen response gene sets (p<0.0001) (**Figure 6A**). Importantly, these reduced activity scores were then significantly increased to a higher level in 4 of the 6 androgen-related gene sets when disease progressed (p<0.05). However, these dynamic changes were not evident in EpiGroup 2 patients with disease progression (**Figure 6B**). We also analyzed dynamics of the activity scores in gene sets of mitotic spindle and p53 pathway since these gene sets showed significant 5hmC increase after 3-month treatment in EpiGroup 3 (**Additional Table S9**). As expected, the activity scores in both gene sets were increased in the EpiGroup 3 when comparing 3-month to baseline (p<0.02, **Additional Figure S4A**). Meanwhile, we also observed a decrease of these activity scores in EpiGroup 2 patients without disease progression (**Additional Figure S4B**). This reduced activity score in EpiGroup 2 was also expected due to relatively higher 5hmC level of the two gene sets in EpiGroups 2 than 3 (**Additional Table S5**). At 24-month, the activity scores of mitotic spindle and p53 pathway remained at the level similar to 3-month.

**Figure 6.**
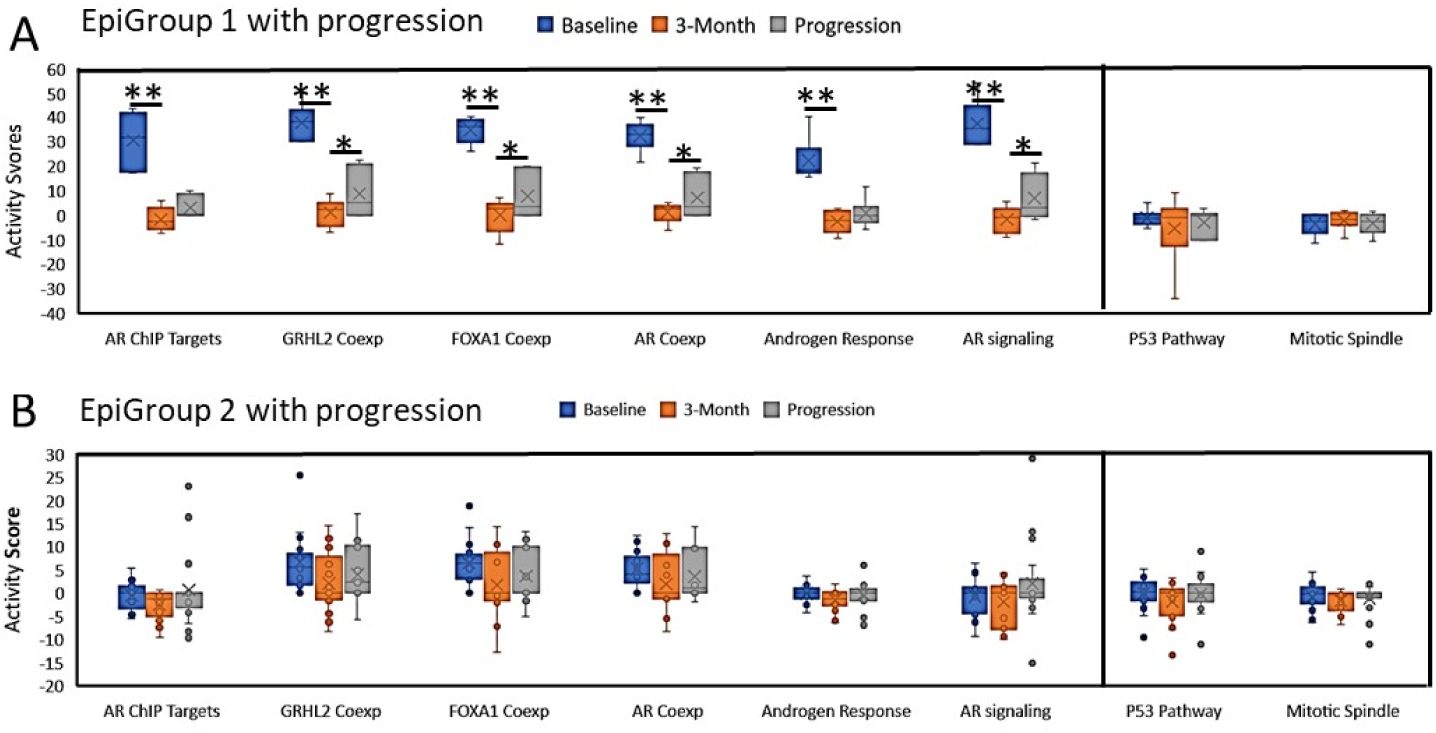
Dynamic changes of activity scores in patients with disease progression during ADT. A. Activity scores in EpiGroup 1 patients were significantly reduced from baseline to 3-month but returned to higher level when disease progressed in all six androgen response gene sets, but not in p53 and mitotic spindle pathways. B. Activity scores in EpiGroup 2 patients with disease progression did not show significant changes in all gene sets tested. Two stars (**) indicate p value<0.0001 when comparing 3-month to baseline. One star (*) represents p value < 0.05 when comparing at progression to 3-month.

## Discussion

Currently, the standard of care for newly diagnosed metastatic castrate-sensitive prostate cancer is to combine LHRH analog and ARSI with or without docetaxel (34). Although there are no head-to-head comparisons, multiple published meta-analysis did not show statistically significant improvement in overall survival with adding docetaxel to LHRH analog and ARSI (triplet therapy) compared to LHRH analog plus ARSI (doublet therapy). Post hoc analysis of the ARASENS trial indicate the overall survival benefit are mainly in high volume but not for low volume metastatic castrate-sensitive prostate cancer(35, 36). Retrospective biomarker studies have reported prognostic biomarkers like mutations in *TP53* or *RB*(37). However, these biomarkers have not been validated in randomized prospective studies. Our study demonstrated that hormone-sensitive prostate cancer patients can be classified into at least 3 epigenomic subgroups with impact on discriminating clinical outcomes. The subgroup with pre-existing androgen signaling activation had shorter time to become castration-resistant. Further validation of this finding will facilitate discovery of more effective biomarker in future clinical trial.

One novel finding from this study was the identification of significant 5hmC enrichment of androgen response gene sets in patients with shorter time to progression on ADT-based therapies. Importantly, this enrichment was present in pre-treatment baseline samples, suggesting potential use for identifying patients who would benefit from upfront treatment intensification. In fact, all patients in this subgroup (N=6) received doublet therapy (4 cases for ADT + docetaxel and 2 cases for ADT + ARSI). Although initial treatment seems effective (as measured by monitoring PSA levels), this group of patients showed rapid disease progression within one year (median PFS=11.73 months). This result suggests inherent resistance to the doublet therapy in this group of patients who featured pre-activation of androgen response genes. Furthermore, activation signal was not limited to hallmark androgen response gene set. We also observed the activation signals from the co-expressed gene sets of *AR*, *FOXA1* and *GRHL2*. Meanwhile, the gene set involved in *AR*-binding targets also showed activation in the subgroup of patients. This observation is consistent with a recent study showing that 5hmC marks the activation of major driver genes in advanced prostate cancer, not only the gene bodies but also downstream target binding sites (16). Based on these observations, we believe that these activity scores not only reflect driver gene activation but also may predict disease progression.

The 5hmC epigenomic changes are highly tissue specific and are more frequently found in genes that drive tissue differentiation and among tissue-specific transcription factors (38, 39). It is more reasonable to speculate the high level of 5hmC in androgen signaling gene sets from prostate cancer origin. However, the current study does not have direct evidence to support that the epigenomic signals were derived from prostate cancer cells only. For example, our study shows significant enrichment of both *FOXA1*- and *GRHL2*-coexpressed genes in patients with disease progression. Using whole blood as RNA source, however, a study found that higher *GRHL2* RNA expression was associated with poor survival in metastatic hormone-sensitive and castration-resistant prostate cancer (40), suggesting that the 5hmC enrichment of *GRHL2* in cfDNA may be derived from blood cells in high risk patients. In contrast, the same study did not observe any association between *FOXA1* RNA expression in blood and poor prognosis. Therefore, the high 5hmC levels in these gene sets are more likely to originate from combination of tumor and nontumor cells. Nevertheless, the gene set activity scores seem effectively separate patients with disease progression from those without.

In addition to predictive value, we also found that the 5hmC-based pharmacodynamic changes in these gene sets may be used to monitor treatment response and disease progression. Our study shows that higher activity scores are significantly reduced in a subgroup of patients with activation in androgen response gene sets. However, this score reduction does not last long before returning to higher levels at disease progression. In addition to androgen signaling, we also observe an increased activity scores of p53 and mitotic spindle pathways in another subgroup patients during the treatment, suggesting activation of the p53-mediated stress response, and cell cycle regulation. These epigenomic changes may reflect the therapeutic effects and influence treatment response and disease progression. Meanwhile, the treatment also induced significant changes in gene sets related to immune response pathways in a subgroup of patients. Clearly, the dynamics of the activity scores during treatment are coincident with treatment response and progression. To our knowledge, this is the first study to demonstrate potential applications of 5hmC-based cfDNA enrichment analysis in predicting treatment response and monitoring disease progression in patients with hormone sensitive prostate cancer, a state that no validated biomarker are currently available for clinical application.

Although the evidence from this study is promising, the current study also has some limitations. First, the study cohort was collected from 2015 to 2019 when the standard care was in transition from ADT to doublet or triplet therapy. Therefore, the study cohort has a significant number of patients (23 of 55) receiving ADT alone. Second, the study cohort is relatively small, which does not allow more detail analysis by further stratifying patients based on their treatment strategies and ethnical groups. Third, this initial discovery was from one cohort only. Lack of an independent validation cohort may lead to cohort-dependent biases. Last, the activation of androgen response gene sets was found in a fraction of patients with early resistance to ADT. Other factors affecting early treatment failure remain unknown. Nevertheless, our study is an initial step to develop an epigenome-based liquid biopsy tool for more effective treatment of the hormone-sensitive prostate cancer.

### Conclusions

Our cfDNA-based 5hmC analysis revealed unique epigenomic profiles in hormone-sensitive prostate cancer patients. 5hmC hypermethylation in androgen response gene sets showed significant association with early resistance to ADT. Activity scores in these gene sets may serve as biomarkers to predict treatment response and monitor disease progression.

## Data Availability

All data produced in the present study are available upon reasonable request to the authors.

## Ethics approval and consent of participation

This study was approved by Institute Review Boards of Medical College of Wisconsin (PRO000023842) and Moffitt Cancer Center (MCC20351)

## Availability of data and materials

The datasets used and/or analyzed during the current study are available from Sequence Read Archive (SRA) submission with accession number xxxxxxx.

## Competing interests

The authors declare that they have no competing interests.

## Authors’ contributions

Conception and design: L.W., D.K., CC.H.

Development of methodology: QX.L., L.W., CC.H.

Acquisition of data: QX.L., L.W., D.K.

Analysis and interpretation of data: QX.L., S.H., YJ.T., JY.H. A.B., M.K., M.P., J.W.

Writing, review, and/or revision of the manuscript: QX.L., M.L., JS.Z., J.Y.P., M.T.N., B.M., M.K., CC.H., L.W.

Administrative, technical, or material support: D.K., E.M.G.

## Acknowledgements

This work is funded by National Institute of Health (R01CA212097 and R01CA250018 to LW). We thank Sequencing Core at the Medical College of Wisconsin for sequencing consultation and support.

## List of abbreviations

ADT: Androgen Deprivation Therapy
FDR: False Discovery Rate
AR: Androgen Receptor
FOXA1: Forkhead Box A1
GRHL2: Grainyhead Like Transcription Factor 2
PSA: Prostate Specific Antigen
ARSI: Androgen Receptor Signaling Inhibitor
cfDNA: cell-free DNA
5hmC: 5-hydroxymethylcytosines
VA: Veteran Affair
NCCN: National Comprehensive Cancer Network
ECOG: Eastern Cooperative Oncology Group
GSEA: Gene Set Enrichment Analysis
ANCOVA: Analysis of Covariance
DMGs: Differentially Methylated Genes
CDS: Coding Sequence
ChIP: Chromatin Immunoprecipitation
LHRH: Luteinizing Hormone-Releasing Hormone

## Figure Legends

**Additional Figure S1. Enrichment efficiency and specificity.** A. PCRs using spike-in templates from non-enriched and enriched DNAs showed specific capture of 5hmC-containing fragments. B. Read count percentages of spike-in DNA in a non-enriched sequencing library. C. Read count percentages of spike-in DNA in an enriched sequencing library.

**Additional Figure S2. Gene set enrichment analysis in differentially methylated genes.** EpiGroup 1 showed significant hypermethylation in the androgen response gene set and significant hypomethylation in immune responses (complement, allograft rejection and inflammation) gene sets when compared to EpiGroup 2 (A, B) or Epigroup 3 (C, D).

**Additional Figure S3. Effect of PSA and ctDNA percentage on clinical outcome and distribution of gene activity at AR locus.** A. Baseline PSA is significantly higher in progressed patients (Y) than non-progressed patients (N). B. ctDNA percentage is significantly higher in progressed patients (Y) than non-progressed patients (N). C. Distribution of read count log2 ratios in EpiGroup 1 (EG1) shows 5hmC enrichment at AR locus but not at AR flanking loci.

**Additional Figure S4. Dynamic changes of activity scores in patients without disease progression during ADT.** A. Activity scores in EpiGroup 3 patients show significant increases in 3 of six androgen signaling gene sets as well as p53 and mitotic spindle gene sets. B. Activity scores in EpiGroup 2 patients were significantly reduced from baseline to 3-month in 2 of six androgen signaling gene sets and mitotic spindle gene set. The star (*) indicates p value <0.05 when comparing 3-month to baseline.

**Additional Table S1.** Detail clinical characteristics of patients

**Additional Table S2.** Total reads, mappable reads and unique reads

**Additional Table S3.** Differentially methylated genes (DMGs) between disease progression and non-progression patients

**Additional Table S4.** Differentially methylated genes (DMGs) among three EpiGroups

**Additional Table S5.** Hallmark gene set enrichment analysis among different EpiGroups

**Additional Table S6.** Analysis of Covariance in patients with different clinical outcomes and different EpiGroups

**Additional Table S7.** Pharmacodynamics of gene 5hmC levels in response to treatment in different EpiGroups

**Additional Table S8.** Gene set enrichment analysis in EpiGroup 1 after 3-month treatment

**Additional Table S9.** Gene set enrichment analysis in EpiGroup 3 after 3-month treatment

## Notes

### Competing Interest Statement

The authors declare no conflict of interest.

### Funding Statement

This study was funded by National Institute of Health (R01CA212097 and R01CA250018 to LW).

### Author Declarations

Institute review boards of Medical College of Wisconsin, Milwaukee VA Medical Center and Moffitt Cancer Center gave ethical approval for this work.

### Summary of Updates

This version was revised to acknowledge significant contribution of two additional authors. This version also provided a detailed methodology in sequence mapping.

## References

1. Bray F, Ferlay J, Soerjomataram I, Siegel RL, Torre LA, Jemal A. Global cancer statistics 2018: GLOBOCAN estimates of incidence and mortality worldwide for 36 cancers in 185 countries. CA Cancer J Clin. 2018;68(6):394–424.

2. Wyatt AW, Azad AA, Volik SV, Annala M, Beja K, McConeghy B, et al. Genomic Alterations in Cell-Free DNA and Enzalutamide Resistance in Castration-Resistant Prostate Cancer. JAMA Oncol. 2016;2(12):1598–606.

3. Maximum androgen blockade in advanced prostate cancer: an overview of the randomised trials. Prostate Cancer Trialists’ Collaborative Group. Lancet. 2000;355(9214):1491–8.

4. Denis L, Murphy GP. Overview of phase III trials on combined androgen treatment in patients with metastatic prostate cancer. Cancer. 1993;72(12 Suppl):3888–95.

5. Sharifi N, Gulley JL, Dahut WL. An update on androgen deprivation therapy for prostate cancer. Endocr Relat Cancer. 2010;17(4):R305–15.

6. Corsini C, Garmo H, Orrason AW, Gedeborg R, Stattin P, Westerberg M. Survival Trend in Individuals With De Novo Metastatic Prostate Cancer After the Introduction of Doublet Therapy. JAMA Netw Open. 2023;6(10):e2336604.

7. Crawford ED, Schellhammer PF, McLeod DG, Moul JW, Higano CS, Shore N, et al. Androgen Receptor Targeted Treatments of Prostate Cancer: 35 Years of Progress with Antiandrogens. J Urol. 2018;200(5):956–66.

8. Kraus LA, Samuel SK, Schmid SM, Dykes DJ, Waud WR, Bissery MC. The mechanism of action of docetaxel (Taxotere) in xenograft models is not limited to bcl-2 phosphorylation. Invest New Drugs. 2003;21(3):259–68.

9. Mistry SJ, Oh WK. New paradigms in microtubule-mediated endocrine signaling in prostate cancer. Mol Cancer Ther. 2013;12(5):555–66.

10. Alix-Panabieres C, Pantel K. Liquid Biopsy: From Discovery to Clinical Application. Cancer Discov. 2021;11(4):858–73.

11. Attard G, Parker C, Eeles RA, Schroder F, Tomlins SA, Tannock I, et al. Prostate cancer. Lancet. 2016;387(10013):70–82.

12. Lone SN, Nisar S, Masoodi T, Singh M, Rizwan A, Hashem S, et al. Liquid biopsy: a step closer to transform diagnosis, prognosis and future of cancer treatments. Mol Cancer. 2022;21(1):79.

13. Nishiyama A, Nakanishi M. Navigating the DNA methylation landscape of cancer. Trends Genet. 2021;37(11):1012–27.

14. Shi DQ, Ali I, Tang J, Yang WC. New Insights into 5hmC DNA Modification: Generation, Distribution and Function. Front Genet. 2017;8:100.

15. Xu Y, Wu F, Tan L, Kong L, Xiong L, Deng J, et al. Genome-wide regulation of 5hmC, 5mC, and gene expression by Tet1 hydroxylase in mouse embryonic stem cells. Mol Cell. 2011;42(4):451–64.

16. Sjostrom M, Zhao SG, Levy S, Zhang M, Ning Y, Shrestha R, et al. The 5-Hydroxymethylcytosine Landscape of Prostate Cancer. Cancer Res. 2022;82(21):3888–902.

17. Song CX, Szulwach KE, Fu Y, Dai Q, Yi C, Li X, et al. Selective chemical labeling reveals the genome-wide distribution of 5-hydroxymethylcytosine. Nat Biotechnol. 2011;29(1):68–72.

18. Scher HI, Halabi S, Tannock I, Morris M, Sternberg CN, Carducci MA, et al. Design and end points of clinical trials for patients with progressive prostate cancer and castrate levels of testosterone: recommendations of the Prostate Cancer Clinical Trials Working Group. J Clin Oncol. 2008;26(7):1148–59.

19. Azad AA, Leibowitz-Amit R, Eigl BJ, Lester R, Wells JC, Murray RN, et al. A retrospective, Canadian multi-center study examining the impact of prior response to abiraterone acetate on efficacy of docetaxel in metastatic castration-resistant prostate cancer. Prostate. 2014;74(15):1544–50.

20. Song CX, Yin S, Ma L, Wheeler A, Chen Y, Zhang Y, et al. 5-Hydroxymethylcytosine signatures in cell-free DNA provide information about tumor types and stages. Cell Res. 2017;27(10):1231–42.

21. Chen S, Zhou Y, Chen Y, Gu J. fastp: an ultra-fast all-in-one FASTQ preprocessor. Bioinformatics. 2018;34(17):i884–i90.

22. Li W, Zhang X, Lu X, You L, Song Y, Luo Z, et al. 5-Hydroxymethylcytosine signatures in circulating cell-free DNA as diagnostic biomarkers for human cancers. Cell Res. 2017;27(10):1243–57.

23. Langmead B, Salzberg SL. Fast gapped-read alignment with Bowtie 2. Nat Methods. 2012;9(4):357–9.

24. Li H, Handsaker B, Wysoker A, Fennell T, Ruan J, Homer N, et al. The Sequence Alignment/Map format and SAMtools. Bioinformatics. 2009;25(16):2078–9.

25. Amemiya HM, Kundaje A, Boyle AP. The ENCODE Blacklist: Identification of Problematic Regions of the Genome. Sci Rep. 2019;9(1):9354.

26. Zhang Y, Liu T, Meyer CA, Eeckhoute J, Johnson DS, Bernstein BE, et al. Model-based analysis of ChIP-Seq (MACS). Genome Biol. 2008;9(9):R137.

27. Patwardhan MN, Wenger CD, Davis ES, Phanstiel DH. Bedtoolsr: An R package for genomic data analysis and manipulation. J Open Source Softw. 2019;4(44).

28. Love MI, Huber W, Anders S. Moderated estimation of fold change and dispersion for RNA-seq data with DESeq2. Genome Biol. 2014;15(12):550.

29. Xie Z, Bailey A, Kuleshov MV, Clarke DJB, Evangelista JE, Jenkins SL, et al. Gene Set Knowledge Discovery with Enrichr. Curr Protoc. 2021;1(3):e90.

30. Beltran H, Prandi D, Mosquera JM, Benelli M, Puca L, Cyrta J, et al. Divergent clonal evolution of castration-resistant neuroendocrine prostate cancer. Nat Med. 2016;22(3):298–305.

31. Adalsteinsson VA, Ha G, Freeman SS, Choudhury AD, Stover DG, Parsons HA, et al. Scalable whole-exome sequencing of cell-free DNA reveals high concordance with metastatic tumors. Nat Commun. 2017;8(1):1324.

32. Du M, Tian Y, Tan W, Wang L, Wang L, Kilari D, et al. Plasma cell-free DNA-based predictors of response to abiraterone acetate/prednisone and prognostic factors in metastatic castration-resistant prostate cancer. Prostate Cancer Prostatic Dis. 2020;23(4):705–13.

33. Liu J, Zhang Y, Li S, Sun F, Wang G, Wei D, et al. Androgen deprivation-induced OPHN1 amplification promotes castration-resistant prostate cancer. Oncol Rep. 2022;47(1).

34. Sanmamed N, Gomez-Rivas J, Buchser D, Montijano M, Gomez-Aparicio MA, Duque-Santana V, et al. Docetaxel Provides Oncological Benefits in the Era of New-Generation Androgen Receptor Inhibitors - or Is Three a Crowd? Clin Genitourin Cancer. 2023.

35. Fizazi K, Foulon S, Carles J, Roubaud G, McDermott R, Flechon A, et al. Abiraterone plus prednisone added to androgen deprivation therapy and docetaxel in de novo metastatic castration-sensitive prostate cancer (PEACE-1): a multicentre, open-label, randomised, phase 3 study with a 2 x 2 factorial design. Lancet. 2022;399(10336):1695–707.

36. Hussain M, Tombal B, Saad F, Fizazi K, Sternberg CN, Crawford ED, et al. Darolutamide Plus Androgen-Deprivation Therapy and Docetaxel in Metastatic Hormone-Sensitive Prostate Cancer by Disease Volume and Risk Subgroups in the Phase III ARASENS Trial. J Clin Oncol. 2023;41(20):3595–607.

37. Kohli M, Tan W, Zheng T, Wang A, Montesinos C, Wong C, et al. Clinical and genomic insights into circulating tumor DNA-based alterations across the spectrum of metastatic hormone-sensitive and castrate-resistant prostate cancer. EBioMedicine. 2020;54:102728.

38. Cui XL, Nie J, Ku J, Dougherty U, West-Szymanski DC, Collin F, et al. A human tissue map of 5-hydroxymethylcytosines exhibits tissue specificity through gene and enhancer modulation. Nat Commun. 2020;11(1):6161.

39. He B, Zhang C, Zhang X, Fan Y, Zeng H, Liu J, et al. Tissue-specific 5-hydroxymethylcytosine landscape of the human genome. Nat Commun. 2021;12(1):4249.

40. Kwan EM, Fettke H, Crumbaker M, Docanto MM, To SQ, Bukczynska P, et al. Whole blood GRHL2 expression as a prognostic biomarker in metastatic hormone-sensitive and castration-resistant prostate cancer. Transl Androl Urol. 2021;10(4):1688–99.

